# Declining use of clean cooking fuels & food security in 2022: Downstream impact of the Russian-Ukrainian war in a Kenyan informal urban settlement

**DOI:** 10.1101/2023.07.09.23292423

**Authors:** Matthew Shupler, James Mwitari, Mark O’Keefe, Federico Lorenzetti, Willah Nabukwangwa, Arthur Gohole, Tash Perros, Emily Nix, Elisa Puzzolo, Daniel Pope, Helen Hoka Osiolo

**Affiliations:** Department of Public Health, Policy and Systems, University of Liverpool, United Kingdom; Kenya Medical Research Institute (KEMRI), Nairobi, Kenya; PayGo Energy Ltd., Nairobi, Kenya; Loughborough University, Loughborough, United Kingdom

**Keywords:** Russian-Ukrainian War, clean cooking, pay-as-you-go, LPG, food security, informal urban settlement

## Abstract

Energy market turmoil due to the Russian-Ukrainian war increased global fuel/food prices. While risks to energy and food security have been suggested, little research has documented impacts for the most vulnerable. During September-October 2022, surveys were administered to 701 households using pay-as-you-go liquefied petroleum gas (PAYG LPG) for clean cooking in an informal settlement in Nairobi, Kenya. Paired t-tests compared PAYG LPG consumption/payment behaviors before (July-October 2021) and during a >15% inflationary food price period (July-October 2022). Three-quarters (74%; %; n=520) of all households and 94% of food insecure households (n=393; 54% of sample) changed their dietary behavior (changed foods cooked, skipped more meals, and/or reduced PAYG LPG consumption) in 2022. Between July-October 2021 and 2022, PAYG LPG prices increased by 16% (214 Kenyan Shilling (KSh)/kg ($1.53USD/kg) to 249 KSh/kg ($1.78USD/kg) and households reduced their monthly PAYG LPG expenditure by 79% (856 KSh ($6.07USD) to 184 KSh ($1.31USD)). Although 97% of participants continued using PAYG LPG in July-October 2022, average consumption declined by two-thirds (0.82 to 0.27 kg/capita/month; difference_(paired-t)_:-0.28 95%CI:[-0.36,-0.21]). Higher food and LPG prices in 2022 led to substantial declines in food security and LPG consumption in an informal urban settlement, highlighting increased obstacles to achieving the Sustainable Development Goals.

**Highlights:**

- 67% reduction (0.82 to 0.27 kg/capita/month) in mean PAYG LPG use between 2021 and 2022
- 97% of homes continued using LPG but monthly fuel expenditure decreased 79% between 2021- 2022
- 56% of households were food insecure
- 94% of food insecure homes changed foods cooked, skipped meals or reduced LPG use in 2022
- One of first studies linking rising food prices with declines in clean energy consumption

*Synopsis:* Using smart meter data, this study illustrates that LPG consumption for cooking dropped by two-thirds and food insecurity increased in an informal urban settlement due to higher food costs in 2022.

## INTRODUCTION

Global food security and access to clean cooking fuels are two of the United Nation’s Sustainable Development Goals (SDG 2 & 7, respectively) for 2030. Food insecurity is defined by The Food and Agricultural Organization (FAO) as having insufficient physical and economic access to safe and nutritious food that meets the dietary needs for an active and healthy life.^1^ In the context of clean cooking, energy insecurity can refer to a lack of access to adequate, affordable, reliable, and clean sources of cooking fuel (e.g. liquefied petroleum gas, electricity) for a healthy and sustainable lifestyle.^2^ Both access to clean household energy and food security are important for reducing poverty (SDG 1), improving health (SDG 3), raising living standards (SDG 10), and reducing gender inequalities (SDG 5).^3^

### Vulnerability in urban sub-Saharan Africa

Sub-Saharan Africa (SSA) has some of the highest global rates of food and energy insecurity; roughly 12% of the region’s population (123 million people) was food insecure in 2022^4^ and 85% of the population (900 million individuals) currently lack access to clean cooking fuels (e.g. liquefied petroleum gas (LPG), electricity).^5^ In SSA, many families live on daily wages and work in the informal economy; families typically spend an average of 60% of their income on food.^4^

Rapid urbanization in SSA is increasing rates of poverty,^6,7^ which is manifested in the form of overcrowded informal urban settlements that often lack adequate housing, infrastructure, sanitation, and health and other social services.^8,9^ The number of SSA households in informal urban settlements has nearly doubled over the last two decades, from 131 million in 2000 to 237 million in 2018.^10^ The increase in informal settlements has contributed to an insufficient supply of household energy for cooking due to the unavailability of wood in urban settings.^11,12^ The urban poor are also more vulnerable to food insecurity than those living in rural settings in SSA due to a lower ability to sustain their family through their land by growing their own food.^13^

The urban poor are also particularly vulnerable to food and energy insecurity during times of crisis.^14–17^ A report by the World Food Program (WFP) found that higher cooking fuel prices can force households to employ various coping strategies, including trading food rations for cooking fuel and skipping meals or undercooking food, as many staple foods in LMICs must be cooked before they are edible.^18^ Additional research has shown that Kenyan households reduced the number of meals, decreased their food variety and consumed more street foods due to increased unemployment and higher prices of staple foods like maize.^14^ Similar declines in food security in 2020 resulted from supply chain disruptions caused by COVID-19 lockdowns and poor harvests.^19,20^ Moreover, 25% of households in a Kenyan informal urban settlement switched away from LPG to polluting cooking fuels that could be bought in smaller amounts (e.g. wood) due to income reductions during COVID-19 lockdowns.^21^

### Measuring the impact of the Russian-Ukrainian war on food and energy security

Following the start of Russian-Ukrainian war in 2022, energy markets became increasingly volatile due to sanctions and oil embargos placed on Russia by Western countries.^22^ food and energy prices rose worldwide due to supply chain disruptions as a result of the war,^23^ which drove up rates of food and energy insecurity.^24–26^ The WFP estimates that up to 47 million and 323 million people may have experienced acute hunger and severe food insecurity, respectively, in 2022.^27^ Studies conducted in high-income countries have shown that individuals reduced their energy consumption due to rising utility bills.^28^

In SSA, where food prices rose by over 15% above 2021 levels in some countries,^29^ the proportion of people suffering from malnutrition or food insecurity increased by over 30% to 123 million, which is 12% of SSA’s population.^4^ While research has shown that the livelihoods of the poor living in LMICs are disproportionately vulnerable to increases in food and energy prices,^30–32^ there is a dearth of data directly measuring the impact of rising costs in 2022 on food and energy security in SSA. Such information can help understand risks to public health during future periods of price instability and inform policies to protect vulnerable households.

Smart meter data from pay-as-you-go liquefied petroleum gas (PAYG LPG) companies operating in East Africa can help fill this data gap. PAYG LPG is a consumer finance mechanism that allows consumers to purchase LPG credits in small increments (via MPesa, a mobile money transfer service available in East Africa).^33^ PAYG LPG companies have typically operated in informal urban settlements to provide clean cooking access to many resource-poor households who would otherwise be unable to afford the upfront costs of LPG equipment.

Through the use of smart meters, the quantity (kilograms) of LPG consumed and length of time using LPG are collected in real-time among customers. This data allows for a longitudinal examination of clean cooking patterns, and has previously been used to demonstrate sustained use of PAYG LPG in Kenya during economic shocks caused by COVID-19 lockdown.^34^

The current study leverages PAYG LPG smart meter data from 2020-2022 alongside cross-sectional surveys and focus group discussions to explore the impact of rising food and energy prices in 2022 on clean cooking consumption and food security in an informal settlement in Nairobi, Kenya.

## METHODS

### Study setting

This study was conducted in Mukuru informal settlement, one of the largest informal settlements in Nairobi. The settlement includes approximately 650 acres of land in an industrial area.^35^ Within Mukuru, our sampling frame included customers of PayGo Energy, a company offering PAYG LPG to residential and small commercial business customers. Founded in 2016, PayGo Energy installs a stainless-steel double burner cookstoves (supplied by *Real Flame* (India)), a gas cylinder, a smart meter and fire safety equipment in customers’ homes. PayGo Energy includes home delivery of cylinder refills and provides customer service support.

### Surveys

From September-October 2022 (7-8 months after the start of the Russian-Ukrainian war in February 2022), 1,122 PayGo Energy customers living in Mukuru informal settlement were invited to participate in a telephonic survey. The telephonic surveys were administered by trained community health volunteers and conducted over telephone via Open Data Kit (ODK) Collect software. Data was encrypted when stored on the ODK Collection application.

A total of 724 individuals (64%) out of the contacted PayGo Energy customers completed the survey. Documented reasons for not completing the survey were being unreachable via phone (n=151; 13%), being uninterested or unwilling to participate (n=100; 9%), being too busy (n=43; 3%), and not receiving a text message notification about the research study (n=22; 2%). Due to potentially large differences in PAYG LPG cooking patterns between residential and commercial customers, an additional 23 customers (2%) that used the PayGo Energy stove for their business were excluded, leaving 701 customers with valid survey data.

The survey asked questions on socioeconomic characteristics, cooking patterns and food consumption. Food security was assessed via the reduced Coping Strategies Index (rCSI),^36,37^ which measures the magnitude of measures taken by households to deal with food insecurity problems.^38^ The rCSI has been validated across many contexts,^37^ including in East Africa.^36^

The rCSI includes five (5) questions about eating less preferred but cheaper foods, reducing the number of meals per day, limiting meal portion sizes at mealtime, prioritizing consumption for children (i.e. limiting adult intake), and borrowing food/money from friends and relatives. Each question asks about frequency in terms of the number of days over the past week. Participants’ responses are weighted and summed (range of scores from 0–56). The rCSI scores were categorized into food secure (0-4), moderately food insecure (4-10), and severely food insecure (≥11) based on previously developed thresholds.^36,38^

### Smart meter data

All customers of PayGo Energy consented to have their smart meter data shared for research purposes when they registered with the company. The anonymized PayGo Energy customer database was stored on a secure, cloud-based server hosted on the Google Cloud Platform and shared securely with University of Liverpool. Anonymized surveys were linked to PayGo Energy’s smart meter data by customers’ account number.

PayGo Energy customers’ LPG consumption and expenditure data included in this analysis spanned from July 2020 (picking up after the period included in a previous paper^34^) through October 2022 (coinciding with the end of survey data collection). In Kenya, significant changes occurred in July of 2021 and 2022 that increased the price of LPG: a 16% VAT was readded on LPG by the Kenyan government in July 2021,^39^ and national price inflation reached 15% in July 2022.^29^

To examine the potential effects of these events on PAYG LPG consumption, the smart meter data was separated into three periods: “pre-VAT on LPG” (July 2020-October 2020), pre inflation period (July 2021-October 2021) and 2022 inflation period (July 2022-October 2022).

To account for seasonal differences in LPG consumption, only data from the months of July-October in each study year (2020-2022) were examined in the main analysis.

The potential impacts of rising food prices on PAYG LPG usage and spending patterns before and after the inflationary period of July-October 2022 were conducted among a subset of 479 households (68% of survey sample) that had been PayGo Energy customers (and therefore had smart meter data) since July 2021 or earlier.

### Statistical analysis

Two-sided paired t-tests were used to examine whether statistically significant ‘within-household’ changes occurred in various PAYG LPG usage (e.g. monthly cooking time, kg LPG/capita/month) and spending metrics (e.g. amount spent per month, number of payments per month) between July-October 2021 and July-October 2022 as a result of inflation. All data that was right-skewed (e.g. kg LPG/capita/month, average payment amount) were log-transformed to meet the assumption of normality prior to the hypothesis testing. For comparison, two-sided paired t-tests were also used to examine potential within-household changes between July-October 2020 and July-October 2021 due to the 16% VAT re-introduced on LPG that took effect nationally in Kenya on 1 July 2021.^39^ All statistical analyses were completed in R version 4.1.1.^40^

### Focus group discussions

From 28 October to 5 November, 2022 (toward the end of the survey data collection period), four focus group discussions (FGDs) were conducted among a random subset of PayGo Energy customers (n=24) to further detail how recent increases in food and energy costs affected their livelihoods. Purposive sampling was used to select customers with equal representation by sex and whether they were generally in charge of food and fuel decisions for their household (yes/no). A total of 9 female decision-makers, 7 female non-decision makers, 4 male decision-makers and 4 male non-decision makers participated.

The FGDs lasted approximately 60-90 minutes and were conducted in Kiswahili by a research coordinator in a central meeting location in Mukuru informal settlement. The FDGs included the following questions related to dietary and cooking patterns in 2022: (1) “what has been your general experience with use of PAYG LPG gas for cooking?” (2) “has your food and PAYG

LPG consumption been affected by the rising cost of food and energy? If so, how?” The FGDs were recorded using a digital recording device with a multi-directional microphone. After transcription and translation into English, key findings from the interviews were identified using thematic analysis in qualitative data analysis software (NVivo version 12). The findings of the interviews are integrated with the quantitative results to provide additional perspective for potential changes in dietary and cooking patterns in 2022.

### Ethical approval

Ethical approval for this study was obtained from the Kenya Medical Research Institute Scientific and Ethics Review Unit, the National Commission for Science, Technology and Innovation (NACOSTI) of Kenya and the University of Liverpool Central Ethics Committee. Informed verbal consent was received from all participants prior to initiating the surveys.

## RESULTS

At the time of survey administration (September-October 2022), less than half (n=308; 44%) of households were food secure, 15% (n=111) were classified as moderately food insecure, and 41% (n=282) were severely food insecure. Nearly all (92%) of food secure customers had received a secondary or university level education, compared with 70% of food insecure individuals (Table 1). Food insecure households were twice as likely to be unemployed (16%) as food secure households (7%) (Table 1). Households with male cooking fuel decision makers were more likely to be food secure (53%) than those with female decision makers (37%) or when the7ecisionn was jointly made by the male and female (37%). The proportion of food secure households cooking exclusively with PAYG LPG or with PAYG LPG and full cylinder LPG (73%) was 11% higher than the percentage of food insecure households cooking exclusively with LPG (62%).

**Table 1.** Characteristics of PayGo Energy customers according to food security status (assessed via the reduced Coping Strategies Index (rCSI))

| Characteristic | Food insecure<br>(n=393) | Food secure<br>(n=308) |
| --- | --- | --- |
| **Highest education level (respondent)** |  |  |
| Primary | 119 (30%) | 26 (8%) |
| Secondary | 191 (49%) | 189 (61%) |
| University/vocational | 83 (21%) | 93 (31%) |
| **Monthly household income** |  |  |
| <10,000 KES | 77 (20%) | 46 (15%) |
| 10,000-17,500 KES | 52 (13%) | 83 (27%) |
| 17,501-30,000 KES | 61 (15%) | 59 (19%) |
| >30,000 KES | 30 (8%) | 39 (13%) |
| Not disclosed | 173 (44%) | 81 (26%) |
| **Occupation** |  |  |
| Salaried job | 32 (8%) | 31 (10%) |
| Business owner | 164 (42%) | 120 (39%) |
| Construction/manufacturing | 36 (9%) | 53 (17%) |
| Informal sector | 92 (23%) | 70 (23%) |
| Unemployed | 61 (16%) | 22 (7%) |
| Other | 8 (2%) | 12 (4%) |
| **Sex of cooking fuel decision maker** |  |  |
| Female | 208 (63%) | 123 (37%) |
| Male | 82 (47%) | 93 (53%) |
| Joint decision | 68 (63%) | 40 (37%) |
| **Marital status** |  |  |
| Single | 79 (20%) | 43 (14%) |
| Married | 298 (76%) | 254 (82%) |
| Divorced/widowed | 16 (4%) | 11 (4%) |
| **Household size** |  |  |
| 1-2 | 50 (13%) | 68 (22%) |
| 3-4 | 178 (45%) | 139 (45%) |
| 5-6 | 132 (34%) | 91 (30%) |
| 7-12 | 33 (8%) | 10 (3%) |
| **% of cooking done using PAYG LPG** |  |  |
| 100% (exclusively used PAYG LPG) | 247 (67%) | 207 (75%) |
| 51-99% | 33 (9%) | 20 (7%) |
| 0-49% | 87 (24%) | 48 (17%) |
| **Cooking fuel types used** |  |  |
| PAYG LPG only | 196 (50%) | 192 (62%) |
| PAYG LPG + full cylinder LPG | 49 (12%) | 35 (11%) |
| PAYG LPG + ethanol | 13 (3%) | 15 (5%) |
| PAYG LPG + charcoal | 65 (17%) | 46 (15%) |
| PAYG LPG + kerosene | 65 (17%) | 19 (6%) |
| PAYG LPG + wood | 5 (1%) | 1 (0%) |

### Changes in dietary and cooking behavior due to rising food and energy prices in 2022

Nearly all (99%; n=692) customers reported an unusual rise in the food price in their community in 2022. Three-quarters (74%; n=520) of participants reported changing their dietary behavior (switched the type of foods cooked, skipped more meals) and/or reduced their PAYG LPG consumption to cope with rising food prices. Nearly two-thirds of households (63%; n=432) elected to eat different foods, half (50%; n=342) had to skip additional meals, and almost one-third (30%; n=201) reported reducing their PAYG LPG consumption due to higher food prices in 2022. Only one customer reported switching their primary cooking fuel from PAYG LPG to a polluting fuel (kerosene) due to the higher fuel price in 2022.

### Changes in dietary and cooking behavior

Both the level of engagement in copies strategies and type of coping strategy undertaken by PayGo Energy customers varied substantially by their food security status (based on their rCSI score). Nearly all (94%; n=371) food insecure households were forced to cope with rising food prices by changing their cooking patterns or reducing their PAYG LPG consumption, compared with half (48%; n=149) of food secure households.

The proportion of food insecure households that changed the foods they cooked and skipped more meals in 2022 (38%; n=151) was six times as high as the percentage of food secure households (6%; n=19) (Figure 1). Moreover, the percentage of food insecure households that engaged in all three coping strategies (changed foods cooked, skipped more meals, reduced PAYG LPG consumption) due to higher food prices in 2022 (24%; n=96) was four times as high as that among food secure households (6%; n=18) (Figure 1).

**Figure 1.**
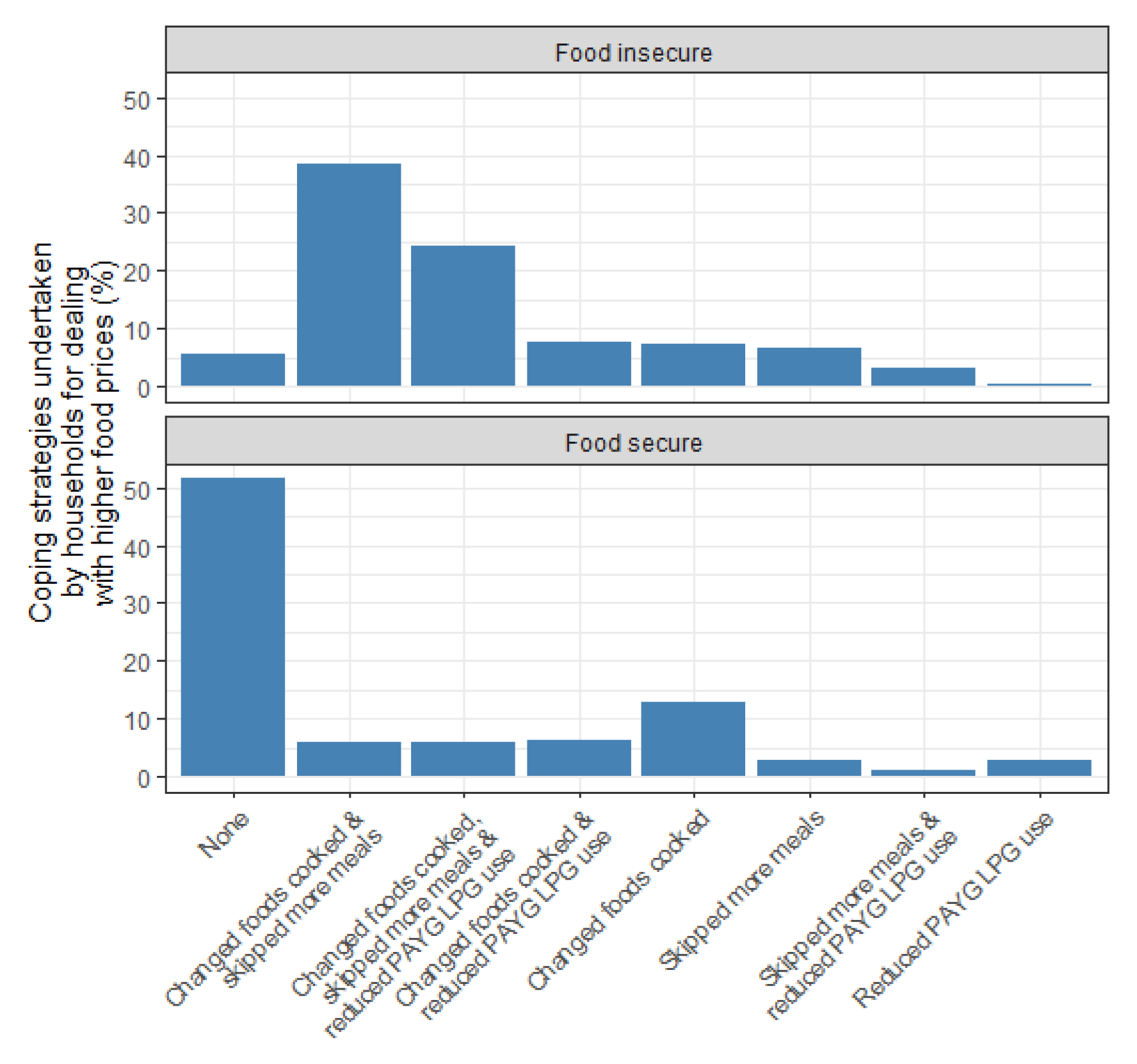
Coping strategies employed by families due to higher food prices in 2022 by food security status.

The proportion of food insecure households that reduced their PAYG LPG consumption to adapt to higher food prices in 2022 (38%; n=146) was twice as high as that among food secure households (19%; n=55). Additionally, 80% (n=226) of households that were classified as severely food insecure skipped more meals in 2022 than 2021, compared with 60% (n=67) of households that were moderately food insecure and 16% (n=49) of food secure households (Supplementary Figure 1). A small proportion (2%; n=11) of households reported that reducing PAYG LPG consumption was the only strategy they undertook to adapt to rising food prices in 2022. The different strategies employed by PayGo Energy customers were highlighted in the FGDs:

> *“I don’t know if it is just on my end, but there has been a high rise in food and fuel [prices], and you have to balance the two. We have had to create a balance by lowering the food and maintaining the gas usage.*” -FGD 2, female respondent
>
> *“You have to budget. Let us say [you have] KSh 200; you cannot spend the whole amount on food but [you must] spare KSh 50 on maybe fuel.”* -FGD 2, female respondent
>
> *“PayGo has come through as a type of fuel during this period but with foodstuff like cooking oil, we have had to reduce portions.”* -FGD 3, male respondent

### Reasons for skipping meals in 2022

The most cited reasons for skipping more meals in 2022 were higher food costs (27%), and lower income (26%). This was also evident in the FGDs:

> *“Our children have a feeding Programme at school which is advantageous so we adults can just skip meals and wait for super to save.”* -FGD 3, male respondent
>
> *“Initially, a 10 kg sack of flour would go for KSh 1850, but right now it goes for KSh 3600, which is double. You have to buy the normal 4 kg [sack]. So it turns out to be hand-to-mouth and also increases the stress level in the house.”* -FGD 4, female respondent

A lower percentage reported lower food availability (10%), a higher cost of PAYG LPG (5%) and drought (4%) as reasons for skipping more meals in 2022.

A higher cost of food was the most common reason reported for skipping more meals in 2022 among food insecure households (43%; n=169), while lower household income was the most frequent cause (9%; n=29) among food secure households (Figure 2). The proportion of food insecure households indicating that higher food costs caused them to skip more meals in 2022 was seven times higher than that among food secure households (6%; n=20). Additionally, 7% (n=29) of food insecure households reporting skipping more meals in 2022 due to higher PAYG LPG prices compared with 1% (n=3) of food secure households (Figure 2).

**Figure 2.**
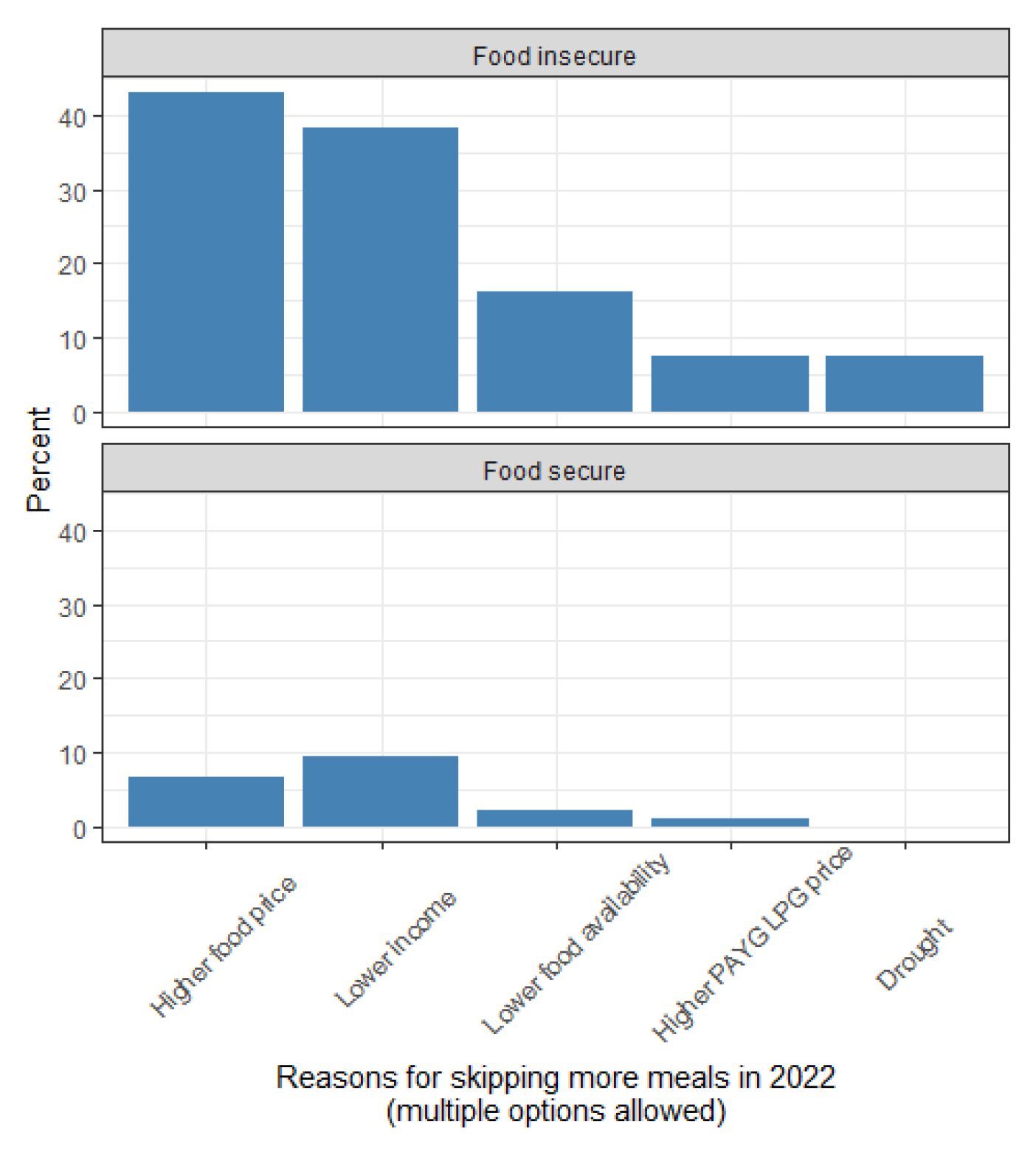
Percent of households reporting specific reasons for skipping more meals in 2022 than 2021 (multiple options allowed)

### Changes in types of food cooked

There were marked fluctuations in the types of foods consumed in 2022, with the degree of dietary changes also varying by food security status. While three quarters of food insecure and secure households consumed more vegetables in 2022, over half of food insecure households reported eating less meat (71%; n=279) and fish (59%; n=233), compared with one quarter of food secure households (20%; n=60% and 25%; n=78, respectively) (Figure 3). The decline in meat consumption was reported in the FGDs:

> *“In my house, we have reduced consumption of luxurious foodstuff like cake and sausages from twice a week to twice a month.”* -FGD 3, male respondent

One quarter of food insecure households (23%; n=90) also reported consuming less ugali, a staple food in Kenya, compared with 10% (n=30) of food secure households. The high cost of maize (the main ingredient in ugali) was frequently referenced in the FDGs conducted among PayGo Energy customers:

> *“You find that 2 kg of maize flour is KSh 250 while you can buy PayGo as low as KSh 100 so food is high. It has affected our finances.”* -FGD 4, female respondent
>
> *“Yes, I have shifted to rice and spaghetti and reduced cooking ugali. Because it takes less time to cook hence saves gas. I can also cook a lot of food during supper and then warm it during lunch hour.”* -FGD 3, male respondent

Additionally, one in five food insecure families and two in five food secure households reported eating more fast food due to rising food prices in 2022 (Figure 3). This was noted in the FGDs:

> *“Initially we would cook chapati and mandazi frequently but decided to reduce and buy already made”* -FGD 3, male respondent

**Figure 3.**
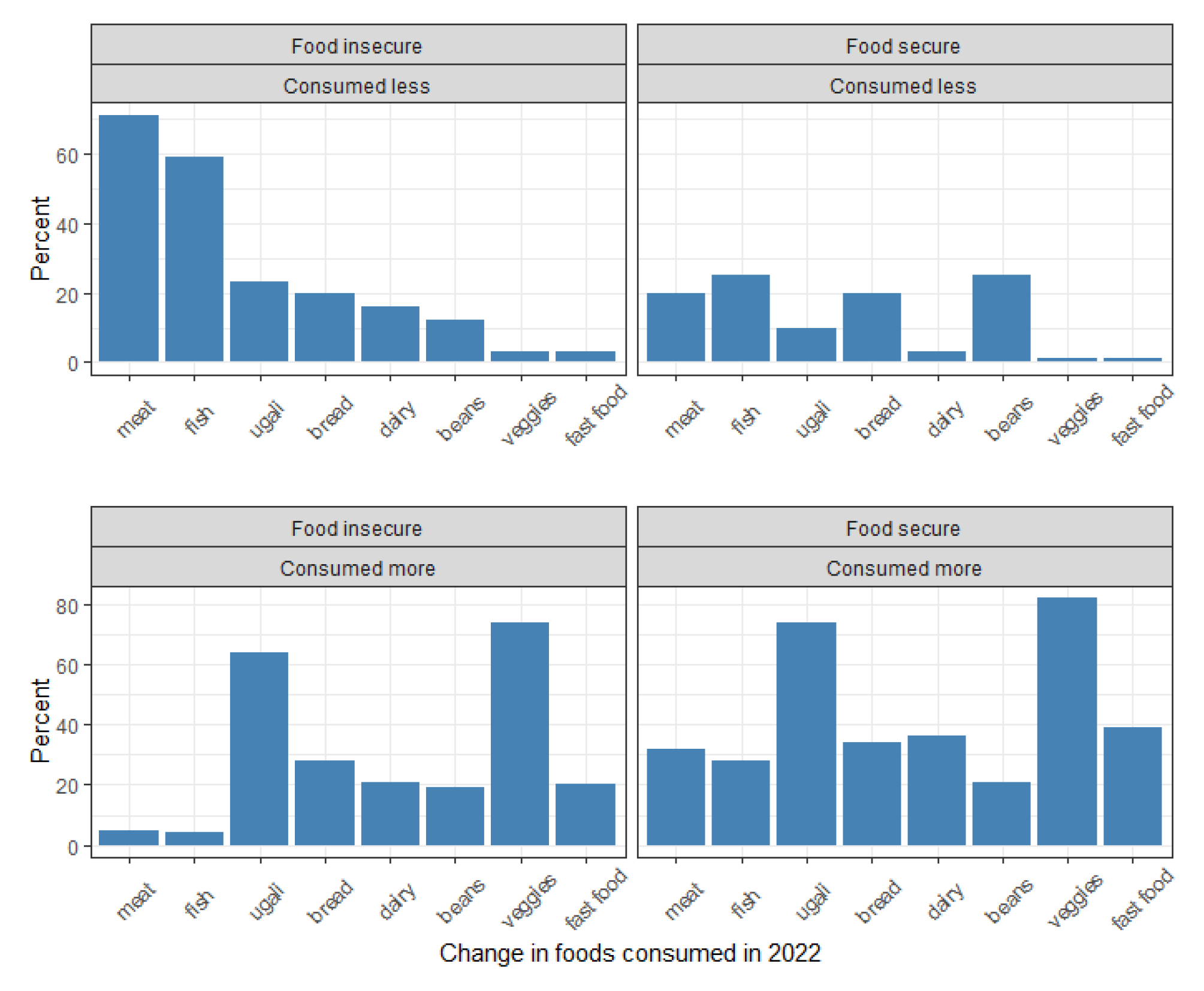
Changes in types of foods cooked as a result of rising food prices in 2022.

### Changes in PAYG LPG consumption due to rising food costs in 2022

During July-October 2021 and July-October 2022, customers cooked with PAYG LPG for an average of 22.1 hours/month, equating to approximately 44 minutes/day (0.73 hours/day).

Individuals used their PAYG LPG stove for an average of 23.8 days/month, corresponding to roughly 6 days per week. During the study period, overall average monthly per capita PAYG LPG consumption was 0.70 kg/capita/month (GSD: 0.78) (Table 3).

**Table 3.**
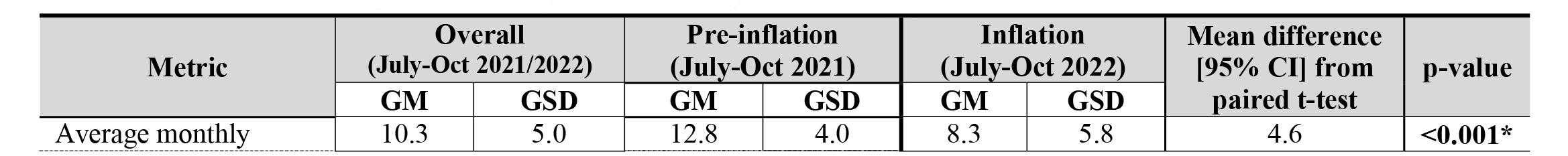
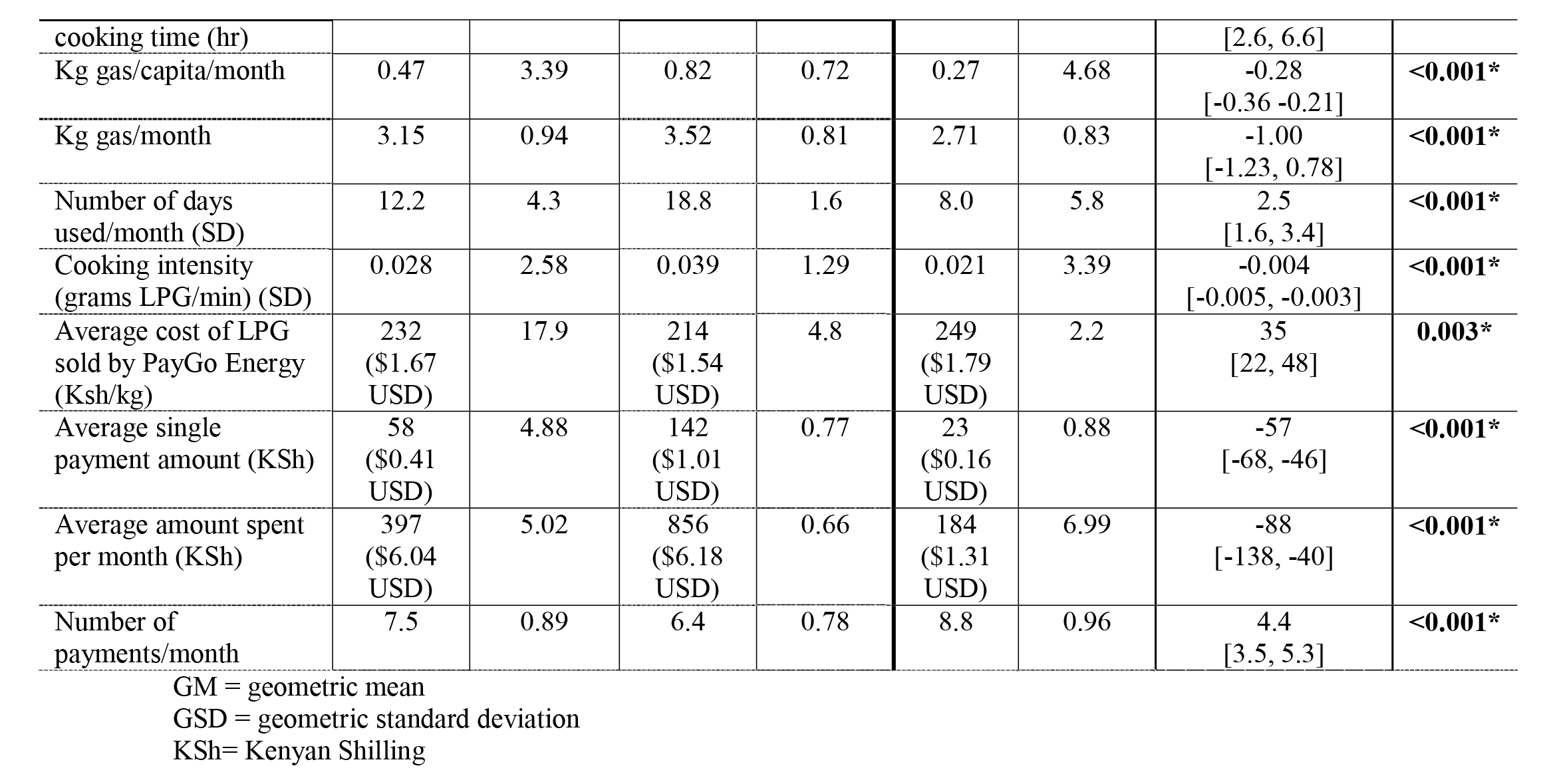
PAYG LPG consumption patterns before and after inflationary food prices due to the Russian-Ukrainian war by food security status (n=297)

According to the smart meter data, 3% (n=15) of households that were customers of PayGo Energy in 2021 completely ceased cooking with PAYG LPG in July 2022. Among the remaining 97% of customers, there was not a significant change in average monthly hours spent cooking with PAYG LPG between July-October 2021 and July-October 2022 (Table 3), during the highest (>15%) months of food price inflation.

Conversely, there was a significant drop in average ‘flame intensity’ (grams of LPG used per minute) while cooking, from 0.044 grams/minute (g/min) in 2021 to 0.041 g/min in 2022 (Table 3). The significant decline in flame intensity contributed to a significant decrease (-0.33 kg/capita/month 95%CI:[-0.42, -0.24]) in average per capita PAYG LPG consumption in July-October 2022 compared with the same months in 2021. Thus, customers’ average consumption during the inflationary period decreased by roughly one-third (35%) from 0.84 kg/capita/month in July-October 2021 to 0.54 kg/capita/month in July-October 2022 (Table 3).

The decline in PAYG LPG consumption was acknowledged by customers in the FGDs:

> *“Sometimes you will eat the same food for lunch that you have cooked for supper [the night before] so you reduce on the fuel consumption.”* -FGD 4, female respondent
>
> *“I also did a practical illustration of the duration of cooking ugali by having [my children] around and concentrating with their phones aside to see the time I took to cook and one of them being a timekeeper. We found out that 5 minutes were enough to cook ugali with maximum concentration and agreed not to exceed 5 minutes to save gas.”* -FGD 4, female respondent

### Changes in PAYG LPG payment patterns due to rising food costs in 2022

PayGo Energy customers reduced their average single payment amount by 36 Kenyan Shilling (KSh) (95%CI:[-45, -27]) ($0.26 USD) from July-October 2021 to July-October 2022 (Table 3). This represents a 40% decrease from the 134 KSh average single payment amount in July-October 2021. However, this decrease was partially offset by a significant increase in the frequency of payments (Figure 3); participants made three more payments (95%CI:[2.2, 3.8]) per month in July-October 2022 compared with July-October 2023, which is a 45% increase from the 6.7 payments/month average in July-October 2021 (Table 3). Nonetheless, customers spent 108 KSh less/month (95%CI:[-161, -55]) on PAYG LPG in July-October 2022 compared with the same months in 2021 (Table 3).

**Figure 3.**
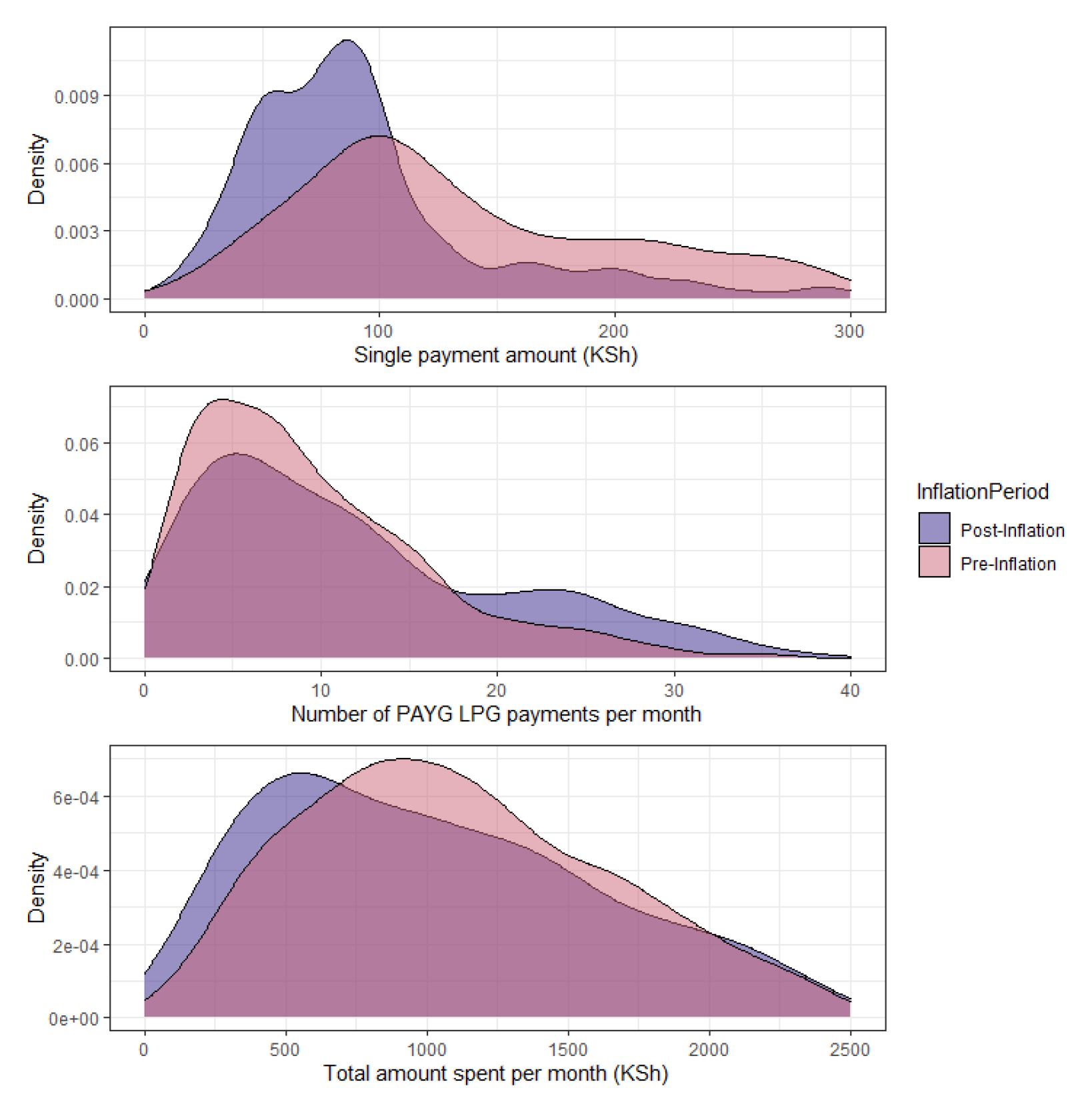
Distribution of MPesa payments for PAYG LPG during pre-inflation (July-October 2021) and post-inflation months (July-October 2022)

### Changes in PAYG LPG consumption due to VAT re-introduction on LPG in 2021

Among the subset of PayGo Energy customers with smart meter data available in 2020-2021 (n=120), average cooking time with PAYG LPG significantly decreased (difference: -4.3 [-7.5, - 1.1] from about 25 to 21 hours/month between July-October 2020 (aftermath of COVID-19 lockdowns) and July-October 2021(VAT was re-added to LPG on 1 July 2021) (Table 4). This was accompanied by a (statistically insignificant) decrease (difference:-0.11 kg/capita/month 95%CI:[-0.30, 0.07]) in average monthly per capita PAYG LPG consumption during the same period (Table 4).

**Table 4.** Pay-as-you-go LPG consumption patterns before and after VAT readded on LPG (July 2021) (n=120)

| Metric | Post COVID-19 lockdown<br>(July–October 2020) |  | Post VAT on LPG<br>(July–October 2021) |  | Mean difference<br>[95% CI] from<br>paired t-test | p-value |
| --- | --- | --- | --- | --- | --- | --- |
|  | GM | GSD | GM | GSD |  |  |
| Average monthly cooking time (hr) | 25.0 | 0.79 | 21.2 | 0.77 | -4.3 [-7.5, -1.1] | **0.008\*** |
| Kg gas/capita/month | 0.91 | 0.76 | 0.79 | 0.75 | -0.11 [-0.30, 0.07] | 0.23 |
| Cooking intensity (grams gas/min) (SD) | 0.043 | 0.15 | 0.044 | 0.27 | -0.0012<br>[-0.0027, 0.0001] | 0.07 |
| Average cost of LPG sold by PayGo Energy (Ksh/kg) | 180<br>(\$1.30 USD) | 0 | 214<br>(\$1.54 USD) | 4.8 | 34<br>[25, 42] | **0.001\*** |
| Average single payment amount | 124 | 0.80 | 104 | 0.84 | -15<br>[-35, 4] | 0.12 |
| Average amount spent per month (KSh) | 695 | 0.74 | 904 | 0.77 | 36<br>[-58, 130] | 0.45 |
| Number of payments/month | 5.5 | 0.91 | 6.8 | 0.96 | 2.2<br>[1.2, 3.2] | **<0.001\*** |

GM = geometric mean
GSD = geometric standard deviation
KSh= Kenyan Shilling

As PayGo Energy passed the added cost of the VAT onto their customers, the PAYG LPG price increased by 34 KSh (180 to 214 KSh/kg) between July-October 2020 and the same months in 2021. While there was a reduction in the average single payment amount between July-October 2020 (124 Ksh) and July-October 2021 (104 KSh), there was no change in total monthly amount spent on PAYG LPG between these two periods (692 vs 698 KSh, respectively) (Table 4). The consistency of total monthly expenditure was due to a statistically significant increase in the number of monthly payments, from an average of 5.5 payments/month in July-October 2020 to 6.8 payments/month in July-October 2021 (Table 4).

## DISCUSSION

By coupling smart meter data with cross-sectional surveys, this study revealed declines in food security and use of PAYG LPG in a Kenyan urban informal settlement due to increased food and energy prices in 2022. Three quarters of PayGo Energy customers were forced to change their cooking and/or dietary patterns in 2022. The coping strategies employed by PayGo Energy customers are consistent with other Kenyan studies; a study conducted in 2016-2017 reported that 81% of households in a community in eastern Kenya adapted to energy shortages by cooking composite meals instead of single meals to reduce the number of cooking sessions per day.^41^

A 16% increase in the cost of PAYG LPG between July-October 2021 and July-October 2022, combined with a 6% decrease in average monthly expenditure on PAYG LPG (Table 3), contributed to the 35% decline (0.84 to 0.54 kg/capita/month) in average per capita monthly consumption between 2021-2022. The smart meter data suggests that, while households continued to use PAYG LPG for cooking for the same number of days each month, they reduced their ‘flame intensity’, which suggests that cooks lowered the flame on their stove or switched from primarily using two burners to one burner on their LPG stove while cooking (Table 3). A potential switch from using two burners to one burner suggests that individuals may have simplified their meals and reduced their food variety.^14^ Other studies conducted in SSA have noted that households can potentially undercook food to conserve fuel.^41,42^

We note that nearly all (97%) of PayGo Energy’s customers continued to cook with PAYG LPG during July-October 2022 despite the higher fuel price, which is consistent with results of a previous study that found 95% of PayGo Energy’s customers maintained their use of PAYG LPG during COVID-19 lockdowns (March-June 2020).^34^ However, the significant decrease in monthly PAYG LPG consumption in 2022 relative to 2021 contrasts with trends from 2020; average monthly per capita PAYG LPG consumption increased during initial COVID-19 lockdowns in April-June 2020 as families were forced to cook more meals at home due to strict stay-at-home orders.^34^

### Comparing VAT re-introduction on LPG to inflation in 2022

The 16% VAT re-added on LPG in 2021 was similar to the ∼15% inflation among food prices that occurred in 2022. Additionally, the 34 KSh increase (180 to 214 KSh) in price of PAYG LPG charged by PayGo Energy in 2021 was similar to the 35 KSh upcharge (214 to 249 KSh) in 2022. However, the reduction in average monthly per capita PAYG LPG consumption between July-October 2020 and the same months in 2021 (-0.11 kg/capita/month) (Table 4) was one-third that of the decline between 2021 and 2022 (-0.33 kg/capita/month) (Table 3). It is possible that the larger increases in food and energy costs are what contributed to greater decreases in PAYG LPG consumption during 2022 relative to 2020 and 2021.^23^ Nonetheless, we note that changes in PAYG LPG payment patterns were similar in 2021 (Table 4) and 2022 (Table 3), with PayGo Energy customers consistently reducing their single payment amount, but increasing their payment frequency. This same pattern emerged among PayGo Energy’s customers during COVID-19 lockdowns in 2020.^34^ This further reinforces the importance of payment flexibility in promoting sustained use of clean cooking during periods of economic instability.^34^ A study conducted during COVID-19 lockdown in 2020 in the same informal settlement found that up to 25% of households cooking with full cylinder LPG (who must purchase a full 6 kg cylinder of LPG upfront) switched to cooking with polluting cooking fuels.^21^

### Disproportionate impact among food insecure households

Compared with approximately half of food secure households, nearly all (94%) food insecure households switched the types of meals they cooked, skipped more meals and/or decreased their PAYG LPG consumption in 2022 relative to 2021. Furthermore, a higher percentage of food insecure PayGo Energy customers (7%) attributed higher PAYG LPG prices as a reason for skipping more meals, compared with food secure households (1%) (Table 2), suggesting that higher energy prices can potentially exacerbate disparities in food security.^43^ This is harmful from a public health standpoint as increased rates of food insecurity can lead to greater deficiencies in essential nutrients.^44^

### Dietary changes

Consumption of meat and fish decreased drastically among the study population (Table 2); this occurred in SSA during other economic crises, including COVID-19 lockdowns^21^ and the 2008 food price crisis.^31^ Some households also reduced their consumption of Kenyan staple foods (e.g. ugali) in 2022 due to greater supply chain disruptions, which were not seen during the COVID- 19 pandemic in 2020.^23^ The price of maize notably rose in 2022 as a result of the Russian-Ukrainian War since the two countries are major maize exporters.^27^

On the other hand, we find that two-thirds of households also increased their consumption of maize (ugali), which has low nutritional value.^45^ Further, we find that one quarter (27%) of households consumed more foods from street vendors (Table 2), which suggests that higher food prices could have made cooking at home more challenging for some households.^14^ This trend can have nutritional implications as street foods tend to be less healthy, due to generally higher quantities of vegetable oil, unhealthy fat, sugar, and ultra-processed food.^46,47^ Previous studies have noted similar increases in consumption of street food during acute crises.^14^

### Health implications

Declines in clean cooking and increased food security following an economic shock have several potential health implications. Firstly, households that reduced their cooking time and skipped additional meals in 2022 are likely to experience increased rates of malnutrition.^48,49^ Moreover, when households change the type of foods cooked during an economic downturn, they often prepare foods that require less time to cook that often have lower nutritional value.^41^ This can further increase the likelihood of malnutrition.

With the exception of one household, none of PayGo Energy’s customers switched their primary cooking fuel from PAYG LPG to a polluting cooking fuel in 2022. This sharply contrasts with the trend among full cylinder LPG users during COVID-19 lockdowns; one-quarter (25%) of these households without access to PAYG LPG switched from full cylinder LPG to polluting fuels due to economic hardships experienced during COVID-19 lockdown.^21^ Thus, access to PAYG LPG may have protected some households from being exposed to higher household air pollution levels that occur when polluting fuels are burned. This is critical from a health perspective as exposure to household air pollution is a risk factor for numerous adverse outcomes, including cardiovascular and respiratory diseases, and is responsible for an estimated 23,000 annual deaths in Kenya.^50^ With an estimated 900 million individuals (∼85% of the population) in SSA currently lacking access to clean cooking fuels,^5^ innovations like PAYG LPG can help poorer households reduce their exposure to household air pollution.

### Strengths and limitations

We objectively evaluated longitudinal changes in cooking patterns by leveraging PAYG LPG smart meter data. We administered surveys and conducted FGDs to help contextualize the findings from the smart meter data analysis. There was a high rate of non-response among surveyed customers (36%). While the smart meter data revealed that the impact of rising food and energy prices due to the Russian-Ukrainian War was similar between survey respondents and survey non-respondents, those that did not participate in the survey had substantially lower levels of PAYG LPG consumption during the study period (0.46 kg gas/month) (Supplementary Table 2) than survey participants (3.15 kg gas/month) (Table 3). This indicates that selection bias may likely be large and that the survey findings are underestimating the true impacts on food insecurity in 2022. For example, the prevalence of severe food insecurity in our study (41%) is less than that obtained from another study also conducted in informal urban settlements in Nairobi (50%).^14^

It is probable that PayGo Energy customers in more precarious financial situations had less time available to conduct a 40-minute survey due to their need to prioritize other income-generating activities. While our survey results may not be generalizable to all PayGo Energy customers and, by extension, all individuals living in the informal settlement, the consistently significant declines in monthly PAYG LPG consumption and payments documented among survey and non-survey participants reinforces the severe, negative impact of rising food and energy costs on individuals living in informal urban settlements in Nairobi.

Although the use of cross-sectional surveys precluded an assessment of a causal association, utilizing two years of real-time, smart meter data allowed for quantification of longer-term cooking patterns prior to, and during, an inflationary period in Kenya. Some survey questions, such as those asking participants if they skipped more meals in 2022 relative to 2021, may be subject to recall bias; however, given the extreme impact of inflation on livelihoods in 2022, we expect recall bias to be minimal. We further used a validated survey tool (rCSI) to obtain an objective measure of food security status at the time of the survey.

Our large sample of PayGo Energy customers facilitated an assessment of longitudinal trends in PAYG LPG consumption in response to economic price shocks in 2021 (VAT on LPG) and 2022 (Russian-Ukrainian War). As the number of PAYG LPG customers continues to increase in SSA, future large-scale studies can leverage smart meter data to understand modifiable determinants of PAYG LPG use to help promote clean cooking.

## CONCLUSIONS

The urban poor in SSA have been negatively impacted by multiple economic shocks in the span of two years (2020-2022). The negative effects of higher food costs were disproportionately experienced by food insecure households, which has likely further widened inequities in food access^23,39^ and increased rates of malnutrition in informal urban settlements in SSA.^48,49^ Although access to PAYG LPG allowed nearly all families in the informal urban settlement to continue their use of clean cooking fuels in 2022 by allowing for incremental fuel payments, rates of LPG consumption dropped by two-thirds from 2021 levels. Thus, the payment flexibility and convenience of PAYG LPG was not sufficient to maintain consistent levels of clean cooking fuel use in 2022. It is therefore critical that governments aim to keep food and fuel prices affordable to buffer low-income households against negative economic shocks. Government policies can include expanding income generation options (which increased access to PAYG LPG has been shown to improve in the same community^51^), increasing the shelf-life of cooked food in areas without access to refrigeration,^41^ leveraging indigenous crops for processed food production,^52^ and providing alternative types of nutritious food products that are cheaper and/or faster to cook.^45,53^

Finally, the expanding availability of smart meter data presents a potential opportunity for real-time tracking of progress towards the Sustainable Development Goals (SDGs), including SDG7 (universal energy access) and SDG 2 (no hunger).^54^ As the African population continues to transition onto digital transaction platforms offered by PAYG LPG and other energy companies, observed trends in energy consumption from smart meter data may be increasingly leveraged to inform results-based financing initiatives and help facilitate targeted social and financial aid programs to the most vulnerable households.

## Data Availability

All data produced in the present study are available upon reasonable request to the authors

## Acknowledgments

We would like to sincerely thank the Community Health Volunteers (CHVs) who conducted the telephonic surveys within Mukuru informal settlement. We would also like to thank the CHVs for openly sharing their experience on the ground with our team during our visit to Nairobi in 2022. We further want to thank the customer care team at PayGo Energy for helping facilitate study logistics. This work was funded by the Sustainable Energy Transitions Initiative early-stage research grant as part of the Environment for Development (EfD) program (Matthew Shupler, Helen Hoka Osiolo). Authors from KEMRI and the University of Liverpool are also supported by funding from the CLEAN-Air(Africa) Global Health Research Unit by the National Institute for Health Research (NIHR) (ref: 17/63/155) using UK aid from the UK government to support global health research.

