## Supplemental Information for "Declining use of clean cooking fuels & food security in 2022: Downstream impact of the Russian-Ukrainian war in a Kenyan informal urban settlement"

Supplementary Table 1. PayGo Energy customers’ reasons for not participating in the telephonic survey

| **Reason** | **N (%)** |
| --- | --- |
| Do not use PayGo Energy stove anymore | 106 (8%) |
| Did not receive equipment | 75 (6%) |
| Not interested | 55 (4%) |
| Too busy | 26 (2%) |
| Did not receive text message alert | 18 (1%) |
| Wrong number | 13 (1%) |

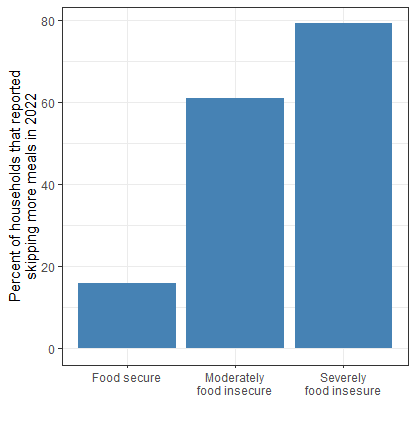

Supplementary Figure 1. Proportion of households reporting skipping more meals in 2022 (relative to 2021) by food security status (derived from reduced Coping Strategies Index (rCSI))

Supplementary Table 2. Pay-as-you-go LPG consumption patterns before and after inflationary food prices due to the Russian-Ukrainian war among households not participating in the survey (n=168)

| **Metric** | **Overall**  **(July-Oct 2021/2022)** | | **Pre-inflation**  **(July-Oct 2021)** | | **Inflation**  **(July-Oct 2022)** | | **Mean difference [95% CI] from paired t-test** | **p-value** |
| --- | --- | --- | --- | --- | --- | --- | --- | --- |
|  | **GM** | **GSD** | **GM** | **GSD** | **GM** | **GSD** |  |  |
| Average monthly cooking time (hr) | 1.57 | 1.55 | 5.96 | 0.79 | 0.15 | 0.80 | -8.39  [-9.57, 7.21] | **<0.001*** |
| Kg gas/month | 0.46 | 7.37 | 2.06 | 3.85 | 0.10 | 9.47 | -1.06  [-1.50, -0.63] | **<0.001*** |
| Number of days used/month (SD) | 20.3 | 10.6 | 17.4 | 5.7 | 23.2 | 5.7 | 5.78  [3.89,7.67] | **<0.001*** |
| Cooking intensity (grams LPG/min) | 0.002 | 7.08 | 0.037 | 1.77 | 0.001 | 9.03 | -0.013  [-0.017, -0.009] | **<0.001*** |
| Average single payment amount (KSh) | 24  ($0.17 USD) | 7.14 | 84  ($0.60 USD) | 4.26 | 7  ($0.05 USD) | 9.00 | -63  [-83, -43] | **<0.001*** |
| Average amount spent per month (KSh) | 408  ($2.90 USD) | 3.54 | 702  ($4.99 USD) | 2.15 | 237  ($1.68 USD) | 4.47 | -73  [-212, 65] | 0.30 |
| Number of payments/month | 7.1 | 0.94 | 6.4 | 0.74 | 7.8 | 0.93 | 4.2  [2.2, 6.1] | **<0.001*** |
